# Impact of Left Common Carotid Cannula Design on Flow Distribution and Cerebral Perfusion Pressure During Bilateral Selective Antegrade Cerebral Perfusion: An Experimental and Computational Study

**DOI:** 10.64898/2026.03.09.26347594

**Authors:** Petter Holmlund, Julia Servin, Axel Vikström, Martha Johannesdottir, Laleh Zarrinkoob, Jan Hellström, Micael Appelblad

**Author notes:** **Corresponding author:** Petter Holmlund, Department of Applied Physics and Electronics, Umeå University, S-901 87 Umeå, Sweden. **This is a preprint that has not been peer-reviewed.**.

## Abstract

**Background:** In aortic arch surgery, bilateral selective antegrade cerebral perfusion (bSACP) maintains cerebral blood flow during circulatory arrest. bSACP is often delivered using a single pump with a Y-connector, dividing the flow. Current practice has veered towards perfusion of the left common carotid artery by cannula and the right subclavian artery or axillary artery by a vascular graft. Under this configuration, inflow distribution may be sensitive to left-sided cannula resistance, particularly in patients with limited collateral circulation, potentially reducing left-hemispheric pressure and flow despite bSACP. We investigated how cannula design influences perfusion pressure and arterial inflow distribution during bSACP.

**Methods:** Four perfusion cannulas with different flow resistances were characterized using bench measurements (40–200 ml/min) and computational fluid dynamics (CFD). The CFD cannula models were then integrated into patient-specific CFD models of the cerebral circulation from three patients with varying collateral circulation/capacity. Both flow- and pressure-controlled pump strategies were simulated.

**Results:** Bench measurements showed substantial variation in flow resistance between the cannulas, which was accurately reproduced by CFD. For the patient-specific analysis, cannula choice affected perfusion through roughly doubled pressure laterality and halved left-side inflow between the most extreme cannulas. Still, perfusion pressure was kept within recommended levels in two subjects but was low in one. Left-side arterial inflow varied between 70-150 ml/min.

**Conclusions:** We isolated the effects of cannula design on cerebral pressure and blood inflow distribution during bSACP, highlighting potential pitfalls in patients with limited collateral circulation.

## A: Introduction

Selective antegrade cerebral perfusion (SACP) is a cardiopulmonary bypass adjunct used during aortic arch surgery to maintain cerebral blood flow during hypothermic circulatory arrest.^1,2^ SACP can be performed either unilaterally (uSACP) or bilaterally (bSACP).^3,4^ uSACP relies on collateral flow through the circle of Willis to supply both hemispheres, whereas bSACP aims to perfuse them more directly. The main rationale for bSACP is to minimize the risk of hypoperfusion in the otherwise indirectly perfused hemisphere^5,6^. bSACP is in clinical practice commonly delivered using a single pump with a Y-connector, dividing flow towards the left and right sides of the head and neck. The most direct variant of bSACP is to insert a cannula inside each common carotid artery (CCA), thereby providing perfusion of both cerebral hemispheres. A common alternative is to perfuse the right side through the right subclavian artery (SA) or axillary artery through a vascular graft, while the left side is perfused via a cannula inserted into the left CCA^7,8^. This method avoids direct cannulation of the right carotid (thereby reducing potential vessel/artery trauma) and allows continuous cerebral perfusion with minimal interruption. A notable drawback of this approach is the risk of asymmetry in hemispheric flow. Because of the smaller diameter of the left CCA, cannulas used on this side are typically smaller than the vascular graft used on the right, which may reduce perfusion pressure and flow in the left hemisphere in patients with limited collateral circulation. This could diminish the intended benefit of bSACP and create a risk of hypoperfusion in the left hemisphere, even under bilateral perfusion. This underscores the importance of selecting the optimal cannula for the left side.

While outer dimensions and insertion techniques for the cannulas are routinely reported in studies utilizing SACP, commonly expressed as French (Fr), the internal geometry of the cannulas and their resulting effects, directly tied to flow resistance, have not been systematically evaluated with respect to perfusion laterality and inflow distribution. Moreover, inflow distribution is rarely reported, with studies instead focusing on total pump flow. A better understanding of how cannula design influences hemispheric perfusion pressure and inflow distribution may therefore aid in achieving appropriate perfusion and in identifying scenarios associated with increased risk.

Mathematical and numerical modeling have increasingly been used to study cardiovascular flow phenomena in cardiopulmonary surgery^9,10^. Computational fluid dynamics (CFD) is one such methodology, enabling the creation of in silico models of different cannula geometries that can be coupled to patient-specific models of the arterial tree, including the circle of Willis, to assess the intraoperative influence of cannula design. We have previously shown that such patient-specific models can reproduce intraoperative/intrasurgical pressure behavior when based on pressure and flow measurements at the cannulation sites.^11^ By incorporating the heart-lung machine tubing, i.e., the Y-connector configuration and cannula, the effects of left-side cannula geometry on perfusion laterality can be explored under clinically realistic pump strategies and collateral conditions.

In this study, we aimed to use bench measurements and CFD to compare commonly used perfusion cannulas and to investigate how the cannula design influences cerebral perfusion pressure and arterial inflow distribution during bSACP. Three aortic arch surgery patients/cases were investigated, two with limited collateral circulation, and one with larger collateral artery size, in order to identify potential risks of hypoperfusion due to internal cannula geometry.

## A: Methods

The method consisted of three stages: First, flow resistance of four arterial perfusion cannulas was characterized using water bench measurements. Second, corresponding computational fluid dynamics (CFD) models of the cannulas were created based on the bench measurements. Third, the resulting CFD cannula models were integrated into patient-specific CFD models of the cerebral circulation for three cases, to quantify their effects on cerebral perfusion pressure and arterial inflow distribution during bSACP. Surgical data from the three cases were used to optimize the anatomical model boundary conditions to reflect intraoperative cerebrovascular conditions.

The study was approved by the Swedish Ethical Review Authority (Dnr: 2022-02770-01) and conducted in accordance with the Declaration of Helsinki. All participants received oral and written information and provided written informed consent.

### B: The cannulas

Four different types of perfusion cannulas commonly used for the left common carotid artery during SACP were tested. The following cannulas were used:

- LeMaitre Distal Perfusion Catheter 2105-15 (LeMaitre Vascular Inc., Burlington, MA, USA), 12 Fr
- SP-GRIPFLOW™ Perfusion Catheter (Fuji Systems Corporation, Tokyo, Japan), 14 Fr
- LivaNova PLC. RCM-14115 Cardioplegia Cannula, Retrograde, ManuallylJInflating Silicone Balloon (LivaNova, Mirandola, Italy), 15 Fr
- True Flow RDB Catheter (European Medical Supplies, S.r.l, Bologna, Italy), 17.4 Fr

Together, these covered a range of flow resistances. The cannulas and connecting parts are displayed in Figure 1.

**Figure 1:**
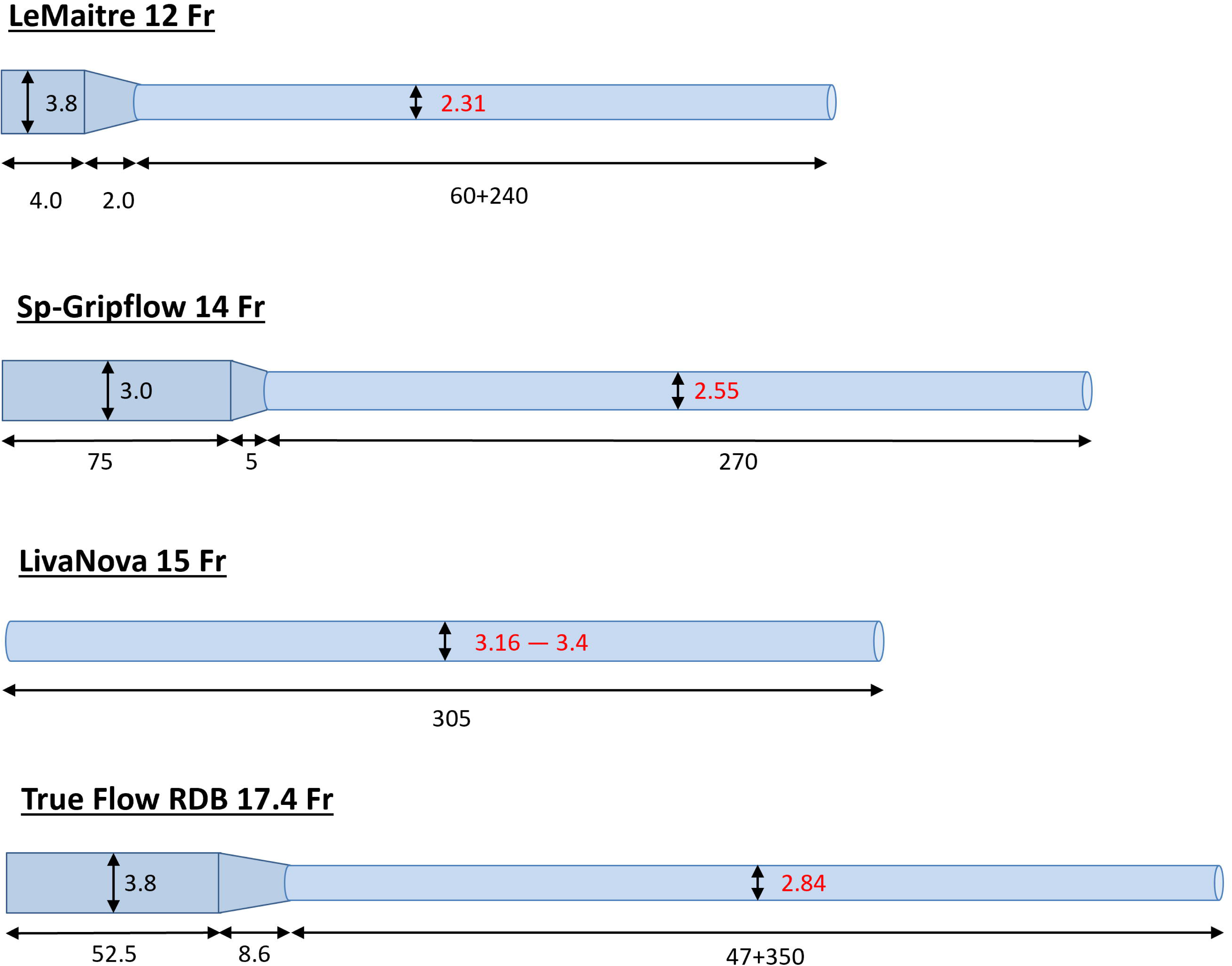
Dimensions of the different cannula designs and their connected tubing. The inner diameters of the actual cannulas had to be determined experimentally (shown in red) as they were not provided by the manufacturers. The LivaNova type cannula has two numbers due to two samples being tested for this type of cannula. All distances are given in mm.

### B: The bench measurements

The bench setup can be seen in Figure 2. Each cannula was connected to a peristaltic pump (Ismatec BVK, Zurich, Switzerland) via silicone tubing (inner radius 4.78 mm, or 3/16 inches) and a rigid plastic connector (Figure 2). The setup allowed steady flow rates between 40 and 175 ml/min.^11^ Pressure was measured immediately upstream of the cannula (inside the connector) using a membrane pressure sensor (PMSET 1DT-XX 1 Safedraw-P, Argon Critical Care Systems, Singapore), positioned at the same hydrostatic level as the cannula to eliminate hydrostatic effects. The cannula outlet was open to the atmosphere. Flow rate was verified by collecting the outflow over 30 seconds in a graduated cylinder. Water temperature (22°C) was used to adjust density and viscosity. For each flow setting, six consecutive pressure and flow measurements were recorded. For the LivaNova cannula type, two samples were tested to also assess intra-design variability.

**Figure 2:**
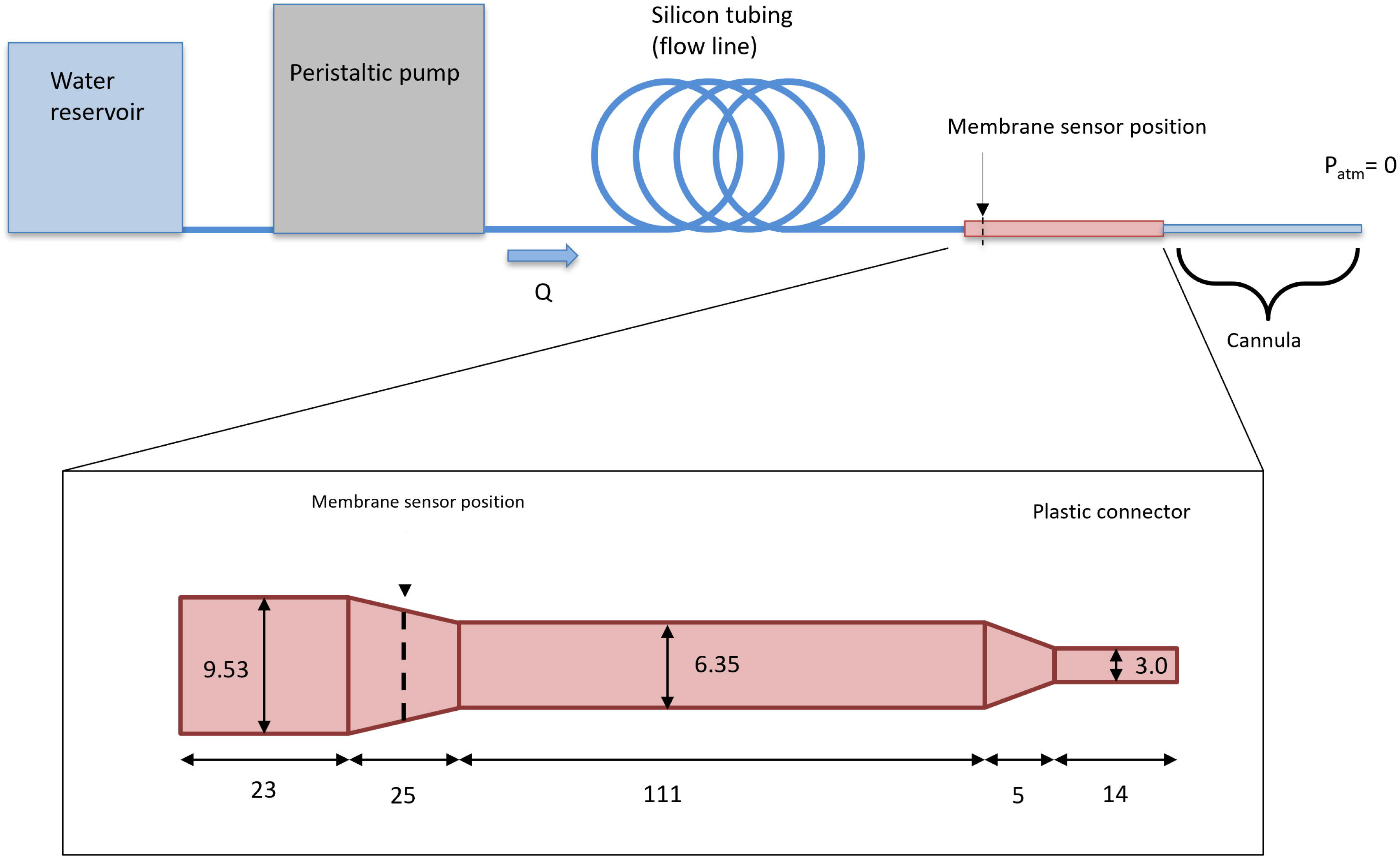
The experimental setup for the bench measurements. Water was pumped through the cannulas using a peristaltic pump providing 6 different flow rates (*Q*). Extended silicon tubing was used to dampen any pulsations in the flow. Pressure was measured using a membrane pressure sensor. The connector tubing (red), attaching the flow line to the cannula, were the same as those used during SACP at Heart Centre, Umeå Sweden. The cannulas (blue) were open to air. Units are given in mm.

### B: CFD cannula model development and validation

CFD simulations were performed in COMSOL Multiphysics® (COMSOL Multiphysics®, version 6.3, https://www.comsol.com, COMSOL AB, Stockholm, Sweden) using the laminar flow physics module, solving the steady-state Navier–Stokes equations for incompressible flow:

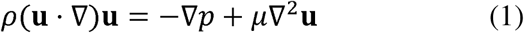

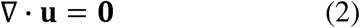

For water at 22 °C, density (*ρ*) and dynamic viscosity (*µ*) were set to 997.8 kg/m^3^ and 0.954 mPa · s, respectively. The inlet boundary was defined as fully developed volumetric flow corresponding to each experimental flow rate, and the outlet pressure was set to atmospheric pressure. The walls were set to no-slip.

Cannula inner radii were estimated using drills of different sizes and refined through sensitivity analyses in COMSOL. All cannulas were subsequently modeled as equivalent cylindrical geometries preserving measured flow resistance to simplify simulations. Mesh convergence was tested (*Table S1)*.

### B: Subject-specific anatomy

Three patient-specific vascular models were created based on preoperative computed tomography angiography (CTA) scans obtained from patients undergoing elective aortic arch surgery at Heart Centre, Umeå University Hospital, Sweden, in 2025^11^ (see Figure 3A for an example). The CTA was performed on the aorto-cervical and major cerebral arteries. The resolution was 0.5 x 0.5 mm^2^ in-plane, and the axial resolution was 0.625 mm. The vascular trees were segmented from the CTA images using Synopsys’ Simpleware (ScanIP P-2019.09, Synopsys, Inc., Mountain View, CA, USA). Images were isotropically interpolated to voxels of 0.3125 mm and to reduce background noise a bilateral filter was applied. The segmented geometries started from the major supra-aortic branches (the brachiocephalic artery, left common carotid artery, and left subaclavian artery), going all the way up to, and including, the circle of Willis. The cerebral outlets of the segmented geometries were the posterior cerebral arteries (PCAs), middle cerebral arteries (MCAs), and distal anterior cerebral arteries (ACA2s), and a lumped outlet for spinal/cerebellar arteries (SpA/CeA). Peripheral outlets included the external carotid arteries (ECAs), left subclavian artery towards the arm, and lumped outlets for the thoracic artery (TA), thyrocervical trunk (TT), and costocervical trunk (CT), one for each side (Figure 3A). The segmentation approach included gradient-based smoothing (local surface correction, search radius: 2 voxels, smoothing radius: 1 voxel), smart mask smoothing (20 iterations), and a small dilation (1 interpolated pixel) to preserve vessel volume.

**Figure 3:**
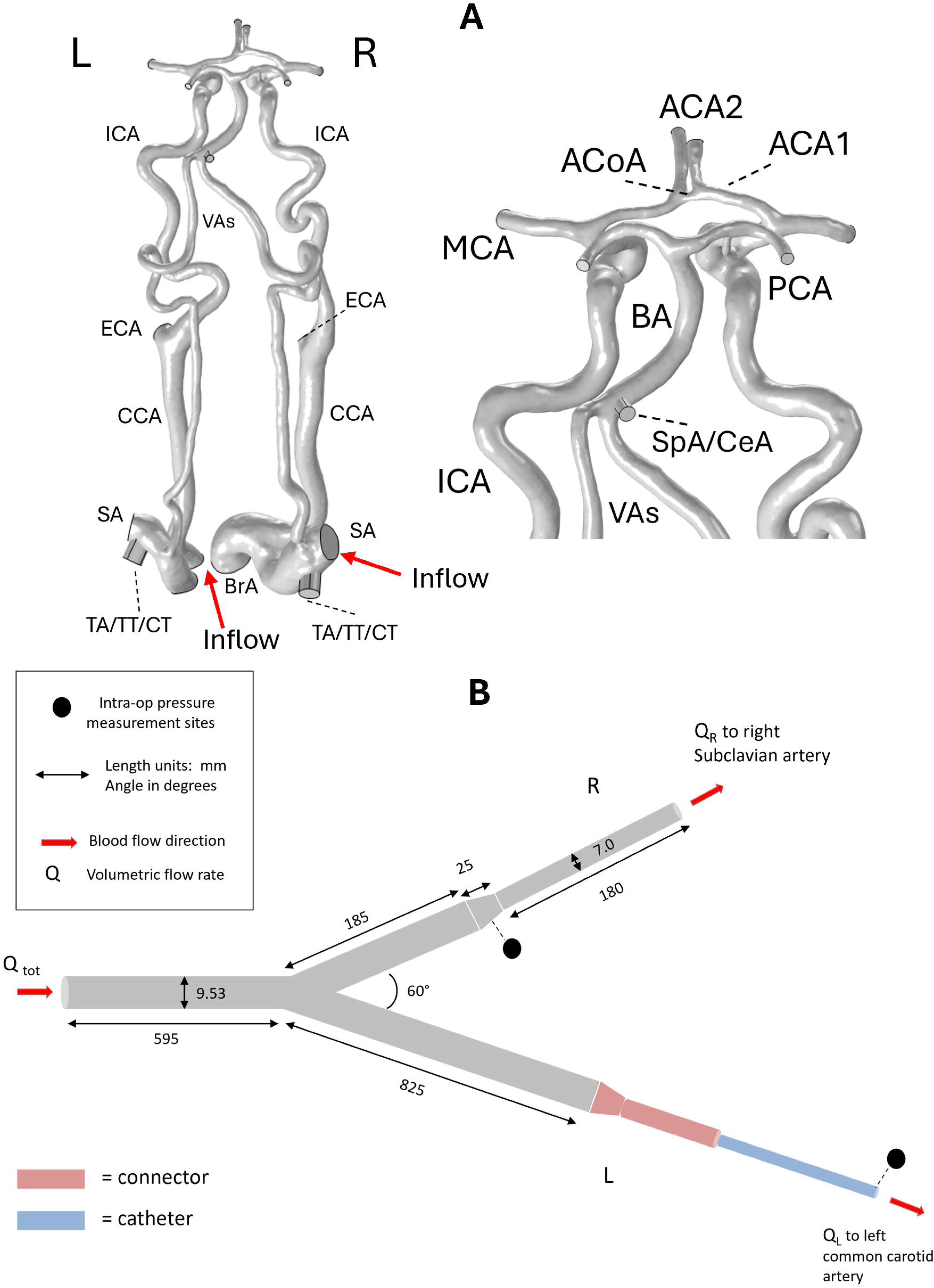
An example arterial tree after segmentation and import to COMSOL, with the right subclavian artery and left common carotid marked as inflow pathways during bSACP (A). A schematic of the Y-tubing connected to the vessel tree (B). Spinal/cerebellar arteries (SpA/CeA),

### B: Surgical setting and in vivo data

The surgery consisted of cardiopulmonary bypass (CPB), using a roller pump (LivaNova S5, Mirandola, Italy) and membrane oxygenation (Affinity Fusion, xx, Holland). The patients were cooled to moderate hypothermia (25 °C in the urinary bladder). CPB was achieved by attaching an 8 mm vascular graft to the right subclavian artery and by cannulating the right atrium with a dual stage cannula. At target temperature circulatory arrest was initiated, the right subclavian, brachiocephalic, and left subclavian arteries were cross-clamped and a 15 Fr Cardioplegia cannula (RCM-14115, LivaNova, Mirandola, Italy) (see Figure 1) was inserted into the left CCA to achieve bSACP for all three subjects. After reaching a steady state with bSACP, uSACP was applied for five minutes. Thereafter, bSACP was re-established and surgery continued. The bSACP data used for this study was collected during five minutes directly following the uSACP. The heart–lung machine was operated in a flow-targeted approach (10 ml//kg/min). The surgical procedure has been described in detail previously.^11^

For achieving bSACP, the graft (right) and the carotid cannula (left) were connected to the heart-lung machine though a Y-connector (Figure 3B) that distributed blood from the heart–lung machine into the right subclavian artery and left common carotid artery, respectively. Relevant measured intraoperative data included:

- total heart-lung machine pump flow
- flow in the left cannula measured by ultrasound (System M, M4; Spectrum Medical, UK)
- pressure at the left cannula tip, pressure just proximal to the graft (and thus the right subclavian artery), and central venous pressure (Monitor: Philips Intellivue MX 800, Andover, MA, USA; Pressure sensors: Codan Xtrans NBSS XL, Forstinning, Germany)
- Hematocrit (Hct) and blood temperature were continuously monitored by the heart-lung machine for viscosity assessment.

Subjects 2 and 3 were selected to represent patients with limited collateral circulation, characterized by smaller anterior communicating vessels compared with subject 1. Subject 1 lacked a posterior communicating artery on the left, and subjects 2-3 had no visible posterior communicating arteries. The subjects’ Willis circles were complete otherwise. Despite similar pump inflow, subjects 2 and 3 differed in mean arterial pressure (MAP), with subject 2 exhibiting substantially higher MAP, reflecting higher total vascular resistance.

### B: Integration of cannulas and patient CFD models

To recreate the surgical setting, the cannula models, graft and Y-connector were merged with each patient’s vascular model to create a complete CFD domain (Figure 4). Mesh sensitivity was analysed for these models as well (see *Table S2*). Blood was approximated as a Newtonian fluid, with density 1060 kg/m^3^ and blood viscosity was calculated based on measured haematocrit and body temperature^12^ (see *Table S3*), resulting in 2.8, 3.3 and 4.3 mPa · s for subject 1, 2 and 3, respectively. An inlet boundary condition was applied at the Y-connector inlet, corresponding to the pump flow setting (inflow or pressure). Cross-clamped arteries were set to no-slip, as were all artery walls. All other arteries were set to outlets. Outflow conditions for all these outlets were defined using a *resistance-type boundary condition*^11,13^ through the equation

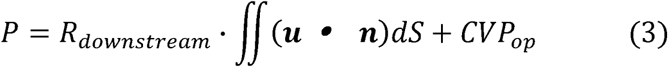

**Figure 4.**
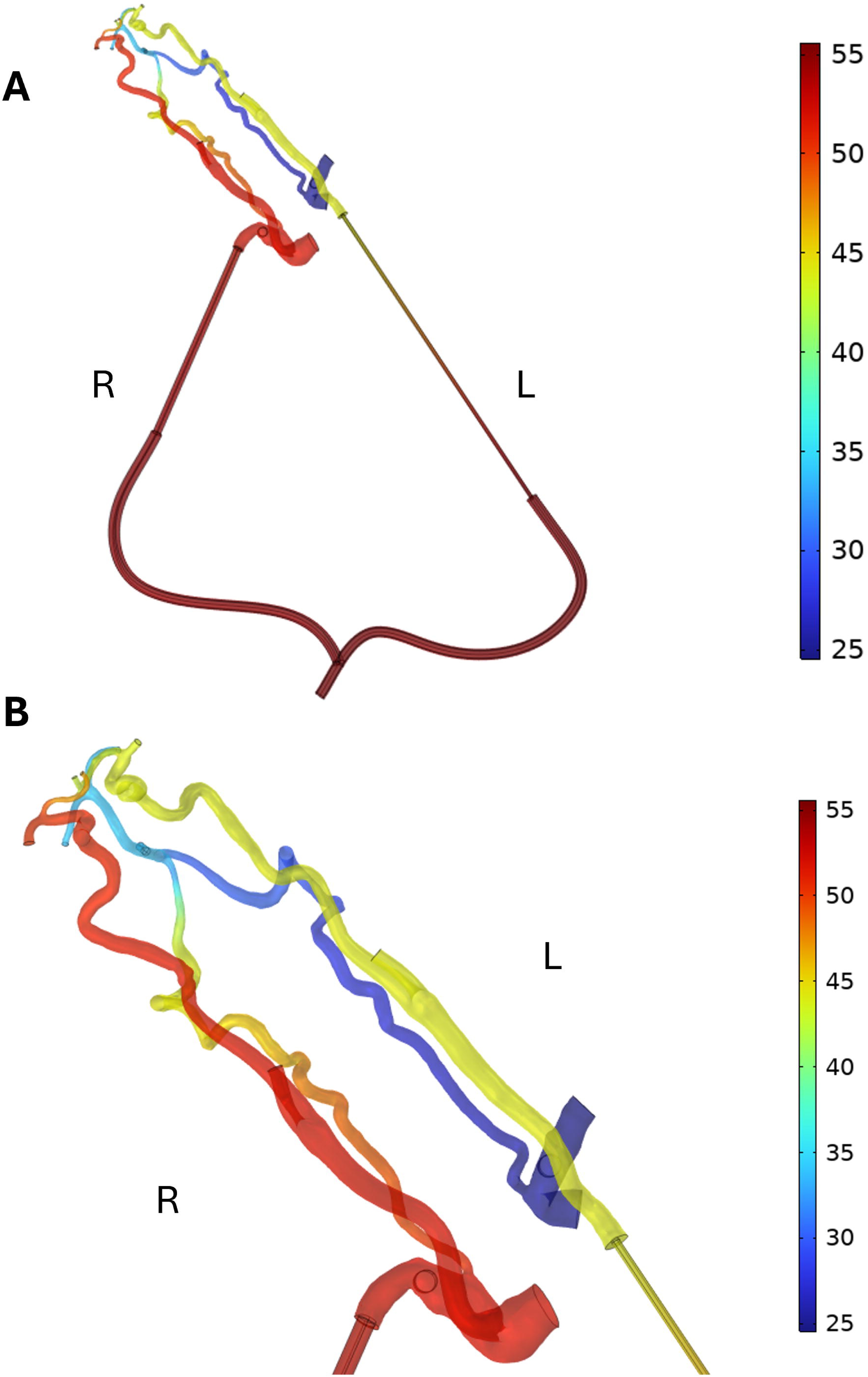
An example image of the connection of the (larger) vascular graft and (thinner) cannula to the anatomical models. The graft and cannula were connected through a Y-connector (A) of tubing 9.53 mm in inner diameter (2.4 mm thickness). The Y-connector has been shortened for illustrative purposes in the figure. The cannula was inserted in the left common carotid artery, and the graft in the right subclavian artery (B). Colour bar represents pressure in mmHg.

where *R_downstream_* represents resistance downstream of the outlet in question, *u* is velocity, *n* the normal vector of the outlet boundary, and *CVP_op_*is the measured central venous pressure during the surgery (4 mmHg, 11 mmHg, and -4 mmHg, for subject 1, 2, and 3, respectively). This boundary condition allows flow to distribute throughout the body based on downstream vascular resistances. The *R_downstream_* values of all vascular territories were calibrated to reproduce the measured intra-operative pressures (within 2 mmHg) during bSACP (see *Table S4*, and *Supplementary materials 1*). This calibration ensured physiologically consistent cerebral boundary conditions. Once calibrated, all downstream resistances were kept identical across simulations and cannula design. This ensured that any changes in perfusion pressure laterality or arterial inflow distribution were attributable to the left-side cannula resistance.

The patient-specific simulations were performed for all four cannula designs under two perfusion control strategies:

1. Flow-targeted, i.e., the pump flows measured intra-operatively.
2. Pressure-targeted, with mean arterial pressure (MAP) set to 50 mmHg.

Both measures are commonly used for guiding these types of surgeries^14,15^. The choice of 50 mmHg comes from being in the mid-to-lower end of the range recommended during cardiopulmonary bypass and SACP^15,16^, and this value is also commonly considered as low during normal circumstances^17^. The primary outcomes for the combined model analyses were:

- Left and right perfusion pressures, denoted *P_L_* and *P_R_*,
- Pressure laterality (difference between right and left side pressures, denoted *dP*), and
- Left cannula flow rate, denoted *Q_L_*, and flow through the graft *Q_R_* (*Q_tot_ - Q_L_ + Q_R_*).

Comparisons were made across all cannulas and patients to assess how cannula resistance influenced these perfusion asymmetry measures.

## A: Results

### B: Bench measurements

There was a clear difference in the pressure drop vs. flow rate relationship for the different types of cannulas (Figure 5), with even some small differences being apparent between the two cannulas of the same type (LivaNova 1 and 2). The cannula with the smallest outer radius (12 Fr, Lemaitre) yielded the highest pressure drops, the Sp-Gripflow (14 Fr) second highest, and the LivaNova (15 Fr) yielded the lowest. The largest cannula in Fr (17.4 Fr, RDB) placed third. CFD simulations could accurately recreate these relationships within the ranges tested (within 1 mmHg per data point) (Figure 5). See *Table S5* for all measurement data. Using blood properties instead of water in the simulations altered absolute pressure levels but did not change the relative resistance differences between cannulas.

**Figure 5:**
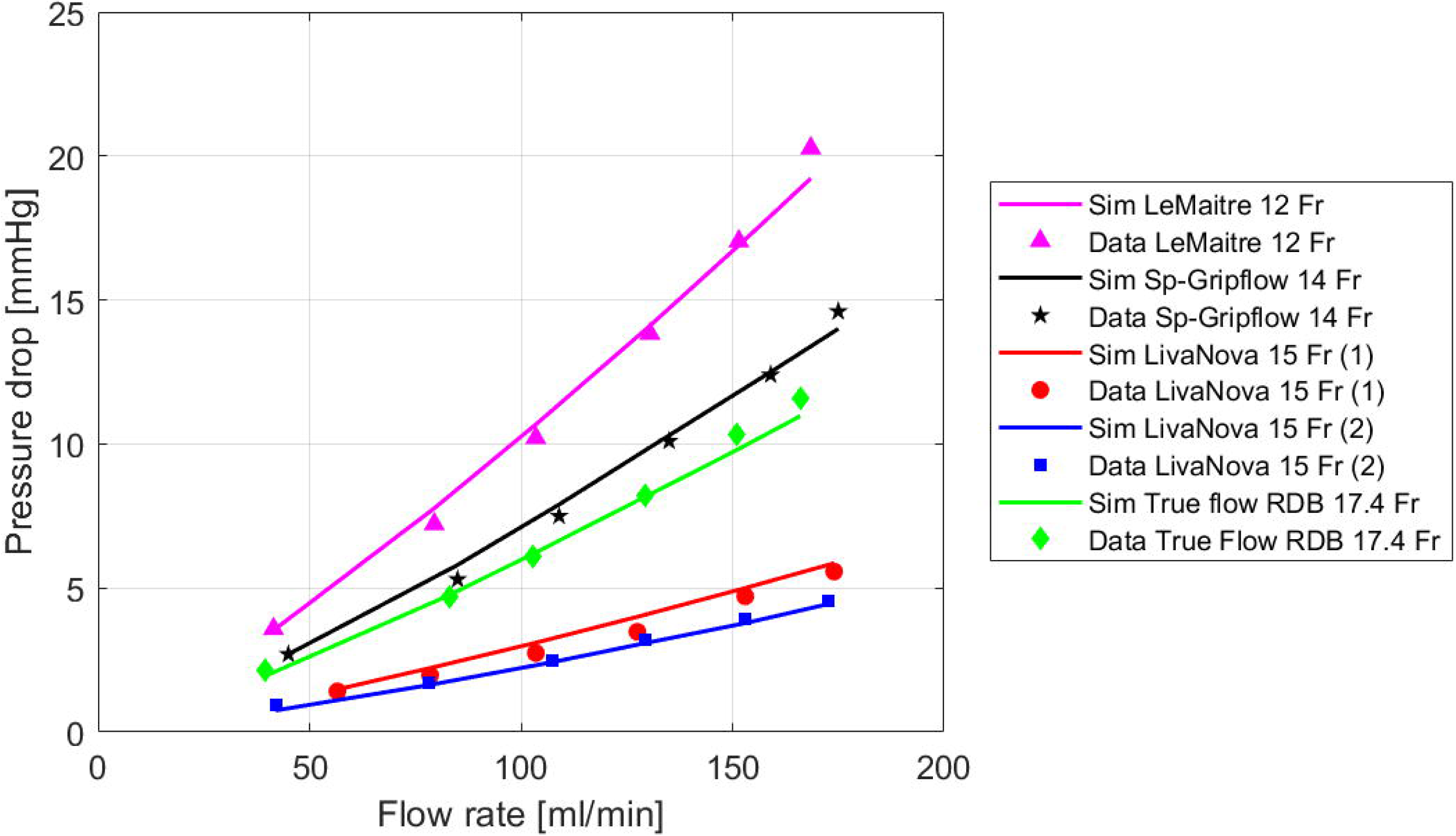
Pressure drop vs. flow rate for the cannula bench measurements. Whole lines correspond to the CFD models, and points correspond to measured data (average of 6 measurements). The Fr number is the outer diameter of the cannulas.

### B: Subject-specific simulations with the different cannulas

Figure 6 (and *Table S6*) shows perfusion pressures and inflow distributions for the different cannulas across the three subjects, under both flow- and pressure-targeted pump strategies. Left-sided pressures were consistently lower than right-sided pressures. For all subjects and control strategies, increasing left-side cannula resistance increased pressure laterality (Δ*P*) and reduced left cannula inflow (*Q_L_*), as total pump flow remained constant. Across the cannulas, pressure laterality more than doubled and *Q_L_* decreased by approximately 50% between the most extreme cannulas.

**Figure 6:**
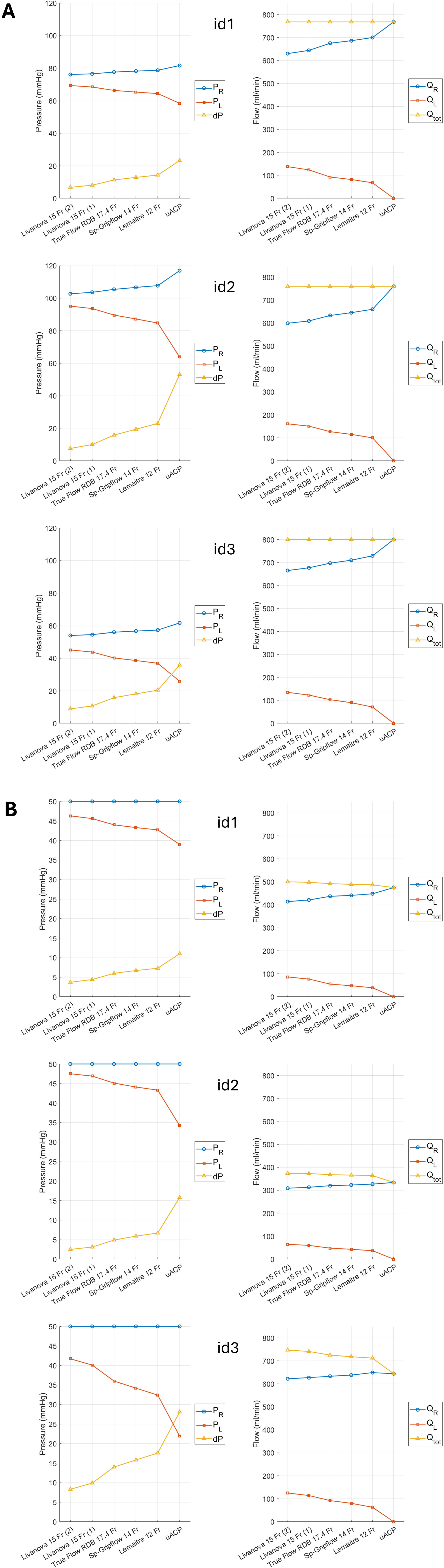
Subject-specific simulations of cerebral perfusion using the different cannulas, as well as uSACP. Perfusion pressure (left column) and inflow distribution (right column) under the flow-targeted pump strategy (A) and pressure-targeted strategy (B). *P_R_* is the pressure in the right subclavian artery, *P_L_* the pressure in the left common carotid artery, *dP* the difference between these two. *Q_R_* is the inflow to the right subclavian artery, *Q_L_* the inflow to the left common carotid, and *Q_to_*_t_ is the total pump flow form the heart-lung machine.

In subject ID3, characterized by low MAP and limited collateral circulation, left-side perfusion pressure fell below 50 mmHg (approached 35 mmHg at the lowest) under flow-targeted control, and fell from roughly 40 mmHg down towards 30 mmHg under pressure-targeted control. The remaining subjects reached pressures down towards 40 mmHg for the pressure-targeted control. Across all configurations, *Q_L_* remained below 150 mL/min.

## A: Discussion

We investigated how left-sided cannula design influences perfusion pressure and inflow distribution during bSACP. Using bench measurements and patient-specific CFD models, we demonstrate substantial differences in flow resistance between commonly used cannulas. These differences may translate into clinically relevant pressure laterality and reduced left-side inflow, particularly in certain patients with limited collateral circulation.

The investigated cannulas represent commonly used perfusion catheter designs from major international manufacturers in contemporary aortic arch surgery. The bench measurements revealed that the highest flow resistance among the investigated cannulas was circa 3.4 times greater than the lowest (Figure 5). Importantly, outer cannula size (Fr) did not reliably predict internal radius or hydraulic resistance. For example, the RDB cannula, despite a relatively large outer diameter, exhibited higher resistance than the LivaNova design due to smaller effective inner dimensions and greater length. While cannula selection in clinical practice depends on several factors — including outer diameter, handling characteristics (which depends on both outer diameter and length), and insertion technique^18^ — internal flow resistance is a distinct property that may warrant greater consideration. As measurements were performed with straight cannulas, intraoperative bending may further increase resistance.

In the patient-specific simulations, increasing left-side cannula resistance consistently reduced left carotid inflow (*Q_L_*) and increased pressure laterality (*dP*) (Figure 6, *Table S6*). Across subjects, *Q_L_* differed by approximately 50% between the most extreme cannulas, despite identical pump settings. Because the left cannula supplies the MCA, ACA, and ECA territories, reduced *Q_L_* increases reliance on collateral crossflow and may predispose to hemispheric hypoperfusion in patients with limited anterior collateral capacity. The intraoperative data suggest narrower anterior circulation in ID2–3 compared with ID1, reflected by greater pressure laterality during uSACP and a larger difference in this laterality between bSACP and uSACP (*Table S4*). Accordingly, simulated *dP* increased more steeply with rising cannula resistance in ID2–3, reaching above 20 mmHg versus 14 mmHg in ID1. In the most vulnerable patient configuration (ID3), left-side perfusion pressure approached commonly cited lower limits.^15,16^ These effects were present under both the flow-targeted and pressure-targeted strategies. Thus, in summary, the intended benefit of bSACP may be attenuated in selected patients by an unfavourable combination of cannula resistance and limited collateral capacity.

The contemporary strategy of perfusing the right subclavian artery and left CCA offers clear advantages compared to bilateral direct CCA cannulation, yet introduces dependency on left-side collateral anatomy and also on cannula resistance. There is much debate as to whether uSACP or bSACP should be the preferred strategy when performing this type of SACP^19–23^ but evidence suggest that bSACP may be preferable during prolonged surgeries.^20,24–26^ Our results indicate, however, that surgical approach in combination with cannula choice can effectively render bSACP semi-unilateral, a nuance that may be negligible in short procedures but could become clinically critical during longer operations, potentially impacting patient safety.

A major strength of this study is the integration of patient-specific intraoperative pressure and flow measurements to establish realistic cerebrovascular conditions. Simulated inflows and pressures matched in vivo measurements for the clinically used cannula (*Table S4* and *Table S6*), with small deviations likely explained by additional resistances in vivo due to cannula bending.

In addition, constant downstream vascular resistances were assumed across simulations, which may be considered a limitation. However, re-scaling/optimizing territorial resistances to uSACP conditions (instead of bSACP conditions) altered *Q_L_* by at most 5 mL/min and *dP* by 2 mmHg, supporting the robustness of the findings. The results are applicable to single-pump perfusion strategies using the right SA and left CCA, and similar configurations.

In conclusion, cannula design influences cerebral perfusion pressure and inflow distribution during bSACP, particularly in patients with limited collateral circulation. Thus cannula-specific resistance could be a factor to consider even during bSACP.

## Author contributions

Conceptualization (PH, MA), Methodology (PH, JS, MA, AV, JH, MJ), Investigation (PH, JS, MA), Analysis (All), Resources (PH, MA, MJ, JH), Data Curation (PH, JS, AV, MA, MR, JH), Writing – Original Draft (PH, MA), Writing – Review & Editing (All).

## Statements and declarations

### Ethical considerations

The study was approved by the Swedish Ethical Review Authority (Dnr: 2022-02770-01) and conducted in accordance with the Declaration of Helsinki.

### Consent to participate

All participants received oral and written information and provided written informed consent.

### Data availability statement

The datasets generated and analysed are not publicly available due to size and imaging data format, but are available from the corresponding author on reasonable request.

