## Supplementary materials 1 for "Impact of Left Common Carotid Cannula Design on Flow Distribution and Cerebral Perfusion Pressure During Bilateral Selective Antegrade Cerebral Perfusion: An Experimental and Computational Study"

*Vessel tree model calibration against in vivo data*

The resistance distribution calibrations of the anatomical models were done by performing bSACP simulations on *only* the anatomical vessel trees of each patient and adjusting the resistances to achieve the in vivo-measured pressures within 2 mmHg (a sort of resistance optimization). Then uSACP simulations were also performed to see that those in vivo pressures also agreed reasonably.

For these simulations, the inlet boundary conditions were: the measured total pump flow rate minus the measured left cannula flow rate ($Q_{r}=Q_{tot}-Q_{l}$) as an inlet condition in the right subclavian artery, and the measured cannula flow $Q_{l}$ in the left CCA (Figure S1). All outlets were based on the resistances from our previous study^11^ and implemented according to Eq. (3). In our previous study, the pre-op resistances were calculated by performing CFD simulations on the CTA data when applying measured flow rates collected from 4D flow MRI sequence (PC-VIPR^27^ measured pre-op in each patient. The 4D flow MRI was performed with a 3T scanner (SIGNATM Premier 3T MRI, GE Healthcare, Milwaukee, WI, USA) using a 48-channel head coil and 30-channel chest-neck coil. Both the arteries of the brain and thorax were scanned, in two separate scans. Scan parameters were, respectively: repetition time 7.2/6.2 ms, echo time 4.95/4.09 ms, velocity encoding 110/150 cm/s (110 used for thorax scan of patient 2), 8° flip angle, 16000 radial projections, acquisition resolution 320x320x320/256x256x256, imaging volume 220x220x220/320x320x320 mm3, isotropic voxel size 0.7x0.7x0.7/1.25x1.25x1.25 mm3, 20 reconstructed cardiac time-frames. The patients’ mean blood pressure (MAP) were monitored using a non-invasive MRI-compatible blood pressure measuring device (CareTaker4, Caretaker Medical, Charlottesville, VA, USA). By using the measured MAP and flow rates, the pressures at the outlets of each artery could be determined ($P_{out}$). From this value, each territory’s resistance could be calculated from Ohm’s law:

$$R_{territory}={(P}_{out}-CVP)/Q_{A} (S1)$$

where $CVP$ is central venous pressure, taken as 4.2 mmHg^28^, and $Q_{A}$ is the outflow through the artery/outlet in question (e.g., the MCA).

Then, the approach to calibrate/optimize the resistances were as follows: The starting values for these resistances were taken from pre-op estimations in each subject, from there, the resistances were multiplied by a scaling factor, one for cerebral vessels (MCA, PCA, ACA2, SpA/CeA), one for the external carotid arteries (ECA) and one for the more peripheral arteries (SA, TA/TT/CT). Second, the scaling factor for the peripheral resistances were lowered. This was done until the left CCA pressure was close to the measured value. Then the cranial resistances were increased, if needed, until the right-side pressure was correct. Finally, the external carotid artery resistance was increased until the correct pressure balance was achieved (within 2 mmHg for the pressure laterality $dP$). The results show that, by this approach, $dP$ and $P_{r}$ and $P_{l}$ could be well recreated when compared to the intra-operative data (*Table S4*). The resulting scaling factors can be seen in Table S7.

Finally, for subject 1, there were more than one set of combinations for the scaling factors that yielded the correct pressures (for subjects 2-3 we only found one combination that worked). To ensure that other scaling factor combinations would not yield different results with respect to our outcome variables ($dP$, $P_{r}, P_{l}$ and $Q_{l}$), a sensitivity test was performed where two different sets of scaling factors were compared (Table S8), showing that the results were not affected.
