## Supplementary material for "Impact of Left Common Carotid Cannula Design on Flow Distribution and Cerebral Perfusion Pressure During Bilateral Selective Antegrade Cerebral Perfusion: An Experimental and Computational Study": Table S1

Table S1. The mesh analysis for one of the LivaNova cannulas. The pressure drop over the cannula vs flow rate was investigated. The **bold** setting was the setting chosen.

|  | Pressure drop [mmHg] | | | |
| --- | --- | --- | --- | --- |
| Flow rate  [ml/min] | Coarse mesh  $\sim100 000$elements | **Normal mesh**  $\boldsymbol{\sim}$**200 000 elements** | Fine mesh  $\sim$500 000 elements | Finer mesh  $\sim$1 million elements |
| 42 | 1.4 | 1.5 | 1.4 | 1.3 |
| 80 | 2.1 | 2.2 | 2.1 | 2.0 |
| 104 | 2.9 | 3.1 | 3.0 | 3.0 |
| 131 | 3.8 | 4.0 | 3.9 | 4.0 |
| 152 | 4.7 | 5.0 | 4.9 | 5.2 |
| 169 | 5.5 | 5.9 | 5.8 | 6.2 |
