## Supplementary material for "Impact of Left Common Carotid Cannula Design on Flow Distribution and Cerebral Perfusion Pressure During Bilateral Selective Antegrade Cerebral Perfusion: An Experimental and Computational Study": Table S2

Table S2. Mesh analysis for the combined Y-coupling + cannula + patient specific anatomy simulations for the three subjects using the LivaNova cannula, flow-controlled pump strategy. The **bold** setting was the setting chosen.

| ID | Mesh Setting | $P_{r}$ | $P_{l}$ | $Q_{left}$ | $dP$ |
| --- | --- | --- | --- | --- | --- |
| 1 | Finer  $\sim$2.7 million elements | 76.4 | 68.6 | 129 | 7.8 |
| 1 | Fine  $\sim$1.5 million elements | 76.3 | 68.6 | 128 | 7.7 |
| 1 | **Normal**  $\boldsymbol{\sim}$**800 000 elements** | 76.5 | 68.4 | 126 | 8.1 |
| 1 | Coarse  $\sim400 000$ elements | 76.7 | 68.5 | 130 | 8.2 |
| 2 | **Finer**  $\boldsymbol{\sim}$**2 million elements** | **50** | **46.9** | **60** | **3.1** |
| 2 | Fine  $\sim$500 000 elements | 50 | 46.5 | 59 | 3.5 |
| 2 | Normal  $\sim$100 000 elements | 50 | 43.6 | 43 | 6.4 |
| 2 | Coarse  $\sim$50 000 elements | 50 | 43.2 | 45 | 6.8 |
| 3 | Finer  $\sim$3 million elements | 54.4 | 44.3 | 127 | 10.1 |
| 3 | Fine  $\sim$1.8 million elements | 54.4 | 44 | 125 | 10.4 |
| 3 | **Normal**  $\sim$**800 000 elements** | 54.5 | 43.7 | 123 | 10.8 |
| 3 | Coarse  $\sim$500 000 elements | 55 | 43.8 | 126 | 11.2 |
