## Supplementary material for "Impact of Left Common Carotid Cannula Design on Flow Distribution and Cerebral Perfusion Pressure During Bilateral Selective Antegrade Cerebral Perfusion: An Experimental and Computational Study": Table S3

Table S3: Viscosity related data from the three subjects during both the uSACP study window and the bSACP study window. Temp stands for body temperature in the urinary bladder, and Hct for hematocrit.

| **ID** | **Viscosity (mPa**$\boldsymbol{\cdot}$**s) *** | **Temp** $\boldsymbol{(^{\circ})}$ | **Hcrt (%)** |
| --- | --- | --- | --- |
| 1 | 2.8 | 23 | 21.3 |
| 2 | 3.3 | 23.4 | 29 |
| 3 | 4.3 | 22.5 | 39.5 |

* Calculated from the temperature and hematocrit.^12^
