## Supplementary material for "Impact of Left Common Carotid Cannula Design on Flow Distribution and Cerebral Perfusion Pressure During Bilateral Selective Antegrade Cerebral Perfusion: An Experimental and Computational Study": Table S4

Table S4: Evaluation of the arterial tree models, i.e., the territorial distal resistance calibration. Simulated pressures versus measured are shown for all three cases. $P_{r}$ for right side pressure (proximal to right subclavian artery) and $P_{l}$ for left side pressure (left CCA pressure), and $dP$ for the difference between the two. Sim indicates simulated in COMSOL.

| **ID** | **SACP type** | $\boldsymbol{dP}$  **[mmHg]** | $\boldsymbol{d}\boldsymbol{P}_{\boldsymbol{sim}}$ **[mmHg]** |  | $\boldsymbol{P}_{\boldsymbol{R}}$  **[mmHg]** | ${\boldsymbol{P}_{\boldsymbol{R}}}_{\boldsymbol{sim}}$  **[mmHg]** |  | $\boldsymbol{P}_{\boldsymbol{L}}$  **[mmHg]** | ${\boldsymbol{P}_{\boldsymbol{L}}}_{\boldsymbol{sim}}$  **[mmHg]** |
| --- | --- | --- | --- | --- | --- | --- | --- | --- | --- |
| 1 | bSACP | 8.9 | 9.8 |  | 76.0 | 76.6 |  | 67.1 | 66.8 |
| 1 | uSACP | 17.9 | 20.7 |  | 68.6 | 75.4 |  | 50.7 | 54.7 |
| 2 | bSACP | 13.0 | 13.3 |  | 103.0 | 104.4 |  | 91.4 | 91.1 |
| 2 | uSACP | 54.2 | 53.1 |  | 107.9 | 116.9 |  | 53.7 | 63.8 |
| 3 | bSACP | 10.0 | 11.0 |  | 52.0 | 54.2 |  | 42.0 | 44.2 |
| 3 | uSACP | 29.1 | 35.9 |  | 64.1 | 63 |  | 35.0 | 27.1 |
