## Supplementary material for "Impact of Left Common Carotid Cannula Design on Flow Distribution and Cerebral Perfusion Pressure During Bilateral Selective Antegrade Cerebral Perfusion: An Experimental and Computational Study": Table S5

Table S5: Measured average flows and pressures for the bench measurements using the four cannula types (mean$\pm$SD). Water at 22 degrees Celsius was used.

| LeMaitre  12 Fr | | Sp-Gripflow  14 Fr | | True Flow RDB  17.4 Fr | |
| --- | --- | --- | --- | --- | --- |
| Flow  [ml/min] | Pressure drop  [mmHg] | Flow  [ml/min] | Pressure drop  [mmHg] | Flow  [ml/min] | Pressure drop  [mmHg] |
| 169$\pm$1.2 | 20.3$\pm$0.1 | 175$\pm$3.1 | 14.6$\pm$0.2 | 166$\pm$2.4 | 11.6$\pm$0.1 |
| 152$\pm$1.6 | 17.0$\pm$0.1 | 159$\pm$1.6 | 12.4$\pm$0.1 | 151$\pm$1.5 | 10.3$\pm$0.1 |
| 131$\pm$1.6 | 13.8$\pm$0.0 | 135$\pm$1.6 | 10.1$\pm$0.2 | 129$\pm2.3$ | 8.2$\pm$0.1 |
| 104$\pm$1.6 | 10.2$\pm$0.1 | 109$\pm$1.5 | 7.5$\pm$0.2 | 103$\pm$1.8 | 6.1$\pm$0.1 |
| 80$\pm$1.6 | 7.2$\pm$0 | 85$\pm$1.5 | 5.3$\pm$0.2 | 83$\pm$1.5 | 4.7$\pm$0.1 |
| 42$\pm$0.5 | 3.6$\pm$0.1 | 45$\pm$0.8 | 2.7$\pm$0.2 | 40$\pm$0.8 | 2.1$\pm$0.0 |

| LivaNova (1)  15 Fr | | LivaNova (2)  15 Fr | |
| --- | --- | --- | --- |
| Flow [ml/min] | Pressure drop  [mmHg] | Flow  [ml/min] | Pressure drop  [mmHg] |
| 174$\pm$1 | 5.6$\pm$0.1 | 173$\pm$1.6 | 4.6$\pm$0.1 |
| 153$\pm$2.7 | 4.7$\pm$0.1 | 153$\pm$2.7 | 3.9$\pm0.1$ |
| 128$\pm$2.5 | 3.5$\pm$0.1 | 130$\pm$0.8 | 3.2$\pm0.1$ |
| 104$\pm$1.6 | 2.7$\pm$0.2 | 107$\pm$0.8 | 2.5$\pm0.1$ |
| 79$\pm$2.9 | 2.0$\pm$0.2 | 78$\pm$0.6 | 1.7$\pm0.1$ |
| 57$\pm$0.7 | 1.4$\pm$0.1 | 42$\pm$0.5 | 0.9$\pm$0.0 |
