## Supplementary material for "Impact of Left Common Carotid Cannula Design on Flow Distribution and Cerebral Perfusion Pressure During Bilateral Selective Antegrade Cerebral Perfusion: An Experimental and Computational Study": Table S6

Table S6: Analysis of perfusion pressure (right side $P_{R}$, left side $P_{L}$, & pressure laterality, i.e., difference between sides $dP$) for the different cannula designs during bSACP in a patient with typical, clearly visible, anterior collateral blood vessels. Inflow distribution is presented through total pump flow ($Q_{tot}$), graft ($Q_{R}$) and cannula flow rate ($Q_{L}$).

| **ID 1**  **Flow-targeted (10ml/kg/mL)** | **Simulated by CFD** | | | | | | | |
| --- | --- | --- | --- | --- | --- | --- | --- | --- |
| **Cannula size** | $\boldsymbol{P}_{\boldsymbol{R}}$ | $\boldsymbol{P}_{\boldsymbol{L}}$ | | $\boldsymbol{dP}$ |  | $\boldsymbol{Q}_{\boldsymbol{R}}$ | $\boldsymbol{Q}_{\boldsymbol{L}}$ | $\boldsymbol{Q}_{\boldsymbol{tot}}$***** |
| LivaNova (2), 15 Fr | 76.1 | 69.3 | | 6.8 |  | 630 | 138 | 768 |
| LivaNova (1), 15 Fr | 76.5 | 68.4 | | 8.1 |  | 644 | 124 | 768 |
| True flow RDB, 17.4 Fr | 77.6 | 66.3 | | 11.3 |  | 675 | 93 | 768 |
| Sp-Gripflow, 14 Fr | 78.2 | 65.3 | | 12.9 |  | 686 | 82 | 768 |
| LeMaitre, 12 Fr | 78.7 | 64.4 | | 14.3 |  | 700 | 68 | 768 |
| uSACP | 81.6 | 58.4 | | 23.2 |  | 768 | 0 | 768 |
| **Pressure-targeted (50 mmHg)** |  | |  | | | | | |
| **Cannula type** | $\boldsymbol{P}_{\boldsymbol{R}}$ | $\boldsymbol{P}_{\boldsymbol{L}}$ | | $\boldsymbol{dP}$ |  | $\boldsymbol{Q}_{\boldsymbol{R}}$ | $\boldsymbol{Q}_{\boldsymbol{L}}$ | $\boldsymbol{Q}_{\boldsymbol{tot}}$ |
| LivaNova (2), 15 Fr | 50 | 46.3 | | 3.7 |  | 414 | 86 | 500 |
| LivaNova (1), 15 Fr | 50 | 45.6 | | 4.4 |  | 421 | 77 | 498 |
| True flow RDB, 17.4 Fr | 50 | 44 | | 6 |  | 437 | 55 | 492 |
| Sp-Gripflow, 14 Fr | 50 | 43.3 | | 6.7 |  | 441 | 48 | 489 |
| LeMaitre, 12 Fr | 50 | 42.7 | | 7.3 |  | 448 | 39 | 487 |
| uSACP | 50 | 39 | | 11 |  | 475 | 0 | 475 |

| **ID 2**  **Flow-targeted (10ml/kg/mL)** |  | |  | | | | | |
| --- | --- | --- | --- | --- | --- | --- | --- | --- |
| **Cannula size** | $\boldsymbol{P}_{\boldsymbol{R}}$ | $\boldsymbol{P}_{\boldsymbol{L}}$ | | $\boldsymbol{dP}$ |  | $\boldsymbol{Q}_{\boldsymbol{R}}$ | $\boldsymbol{Q}_{\boldsymbol{L}}$ | $\boldsymbol{Q}_{\boldsymbol{tot}}$***** |
| LivaNova (2), 15 Fr | 102.7 | 95.1 | | 7.6 |  | 599 | 161 | 760 |
| LivaNova (1), 15 Fr | 103.6 | 93.6 | | 10 |  | 609 | 151 | 760 |
| True flow RDB, 17.4 Fr | 105.4 | 89.6 | | 15.8 |  | 633 | 127 | 760 |
| Sp-Gripflow, 14 Fr | 106.6 | 87.2 | | 19.4 |  | 645 | 115 | 760 |
| LeMaitre, 12 Fr | 107.7 | 84.7 | | 23 |  | 660 | 100 | 760 |
| uSACP | 116.9 | 63.8 | | 53.1 |  | 760 | 0 | 760 |

| **Pressure-targeted (50 mmHg)** |  | |  | | | | | |
| --- | --- | --- | --- | --- | --- | --- | --- | --- |
| **Cannula type** | $\boldsymbol{P}_{\boldsymbol{R}}$ | $\boldsymbol{P}_{\boldsymbol{L}}$ | | $\boldsymbol{dP}$ |  | $\boldsymbol{Q}_{\boldsymbol{R}}$ | $\boldsymbol{Q}_{\boldsymbol{L}}$ | $\boldsymbol{Q}_{\boldsymbol{to}\boldsymbol{t}}$ |
| LivaNova (2), 15 Fr | 50 | 47.5 | | 2.5 |  | 309 | 65 | 374 |
| LivaNova (1), 15 Fr | 50 | 46.5 | | 3.5 |  | 314 | 59 | 373 |
| True flow RDB, 17.4 Fr | 50 | 45.1 | | 4.9 |  | 320 | 48 | 368 |
| Sp-Gripflow, 14 Fr | 50 | 44.1 | | 5.9 |  | 323 | 43 | 366 |
| LeMaitre, 12 Fr | 50 | 43.3 | | 6.7 |  | 327 | 37 | 364 |
| uSACP | 50 | 34.2 | | 15.8 |  | 334 | 0 | 334 |

| **ID 3**  **Flow-targeted** |  | | | | | | |
| --- | --- | --- | --- | --- | --- | --- | --- |
| **Cannula size** | $\boldsymbol{P}_{\boldsymbol{R}}$ | $\boldsymbol{P}_{\boldsymbol{L}}$ | $\boldsymbol{dP}$ |  | $\boldsymbol{Q}_{\boldsymbol{R}}$ | $\boldsymbol{Q}_{\boldsymbol{L}}$ | $\boldsymbol{Q}_{\boldsymbol{tot}}$***** |
| LivaNova (2), 15 Fr | 54 | 45.1 | 8.9 |  | 665 | 135 | 800 |
| LivaNova (1), 15 Fr | 54.5 | 43.7 | 10.7 |  | 677 | 123 | 800 |
| True flow RDB, 17.4 Fr | 56 | 40.2 | 15.8 |  | 697 | 103 | 800 |
| Sp-Gripflow, 14 Fr | 56.7 | 38.6 | 18.1 |  | 710 | 90 | 800 |
| LeMaitre, 12 Fr | 57.3 | 36.8 | 20.5 |  | 729 | 71 | 800 |
| uSACP | 61.7 | 25.9 | 35.8 |  | 800 | 0 | 800 |

| **Pressure-targeted (50 mmHg)** |  | |  | | | | | |
| --- | --- | --- | --- | --- | --- | --- | --- | --- |
| **Cannula type** | $\boldsymbol{P}_{\boldsymbol{R}}$ | $\boldsymbol{P}_{\boldsymbol{L}}$ | | $\boldsymbol{dP}$ |  | $\boldsymbol{Q}_{\boldsymbol{R}}$ | $\boldsymbol{Q}_{\boldsymbol{L}}$ | $\boldsymbol{Q}_{\boldsymbol{tot}}$ |
| LivaNova (2), 15 Fr | 50 | 41.7 | | 8.3 |  | 622 | 125 | 747 |
| LivaNova (1), 15 Fr | 50 | 40.1 | | 9.9 |  | 627 | 114 | 741 |
| True flow RDB, 17.4 Fr | 50 | 36 | | 14 |  | 633 | 92 | 725 |
| Sp-Gripflow, 14 Fr | 50 | 34.2 | | 15.8 |  | 638 | 80 | 718 |
| LeMaitre, 12 Fr | 50 | 32.4 | | 17.6 |  | 649 | 63 | 712 |
| uSACP | 50 | 21.9 | | 28.1 |  | 644 | 0 | 644 |

***** Corresponds to the measured $Q_{tot}$ during the actual surgery. For the pressure-targeted scheme, $Q_{tot}$ was simulated.
