## Supplementary material for "Impact of Left Common Carotid Cannula Design on Flow Distribution and Cerebral Perfusion Pressure During Bilateral Selective Antegrade Cerebral Perfusion: An Experimental and Computational Study": Table S7

Table S7. The scaling factors used for each patient to achieve the resistances that achieve the measured pressures at surgery.

| Scaling factor | ID1 | ID2 | ID3 |
| --- | --- | --- | --- |
| Peripheral scaling | 0.85 | 0.42 | 0.45 |
| Cranial scaling | 0.96 | 1.02 | 1.23 |
| ECA scaling | 0.96 | 1.02 | 1.23 |
