## Supplementary material for "Impact of Left Common Carotid Cannula Design on Flow Distribution and Cerebral Perfusion Pressure During Bilateral Selective Antegrade Cerebral Perfusion: An Experimental and Computational Study": Table S8

Table S8. For ID1, two sets of parameters were used to test any differences in the pressures calculated. Highly similar results were achieved for both configurations.

| **Original setting** | Peripheral scaling | Cranial  scaling | ECA scaling | $P_{r}$ | $P_{l}$ | $dP$ |
| --- | --- | --- | --- | --- | --- | --- |
| bSACP | 0.85 | 0.96 | 0.96 | 76.6 | 66.8 | 9.8 |
| uSACP | 0.85 | 0.96 | 0.96 | 75.4 | 54.7 | 20.7 |
| **Alternate setting** | Peripheral scaling | Cranial  scaling | ECA scaling | $P_{r}$ | $P_{l}$ | $dP$ |
| bSACP | 0.89 | 1.01 | 0.89 | 77.4 | 67.3 | 10.1 |
| uSACP | 0.89 | 1.01 | 0.89 | 75.3 | 55.1 | 20.2 |
